# Genomic analyses of human adenoviruses unravel novel recombinant genotypes associated with severe infections in pediatric patients

**DOI:** 10.1101/2021.08.01.21261368

**Authors:** Joyce Odeke Akello, Richard Kamgang, Maria Teresa Barbani, Franziska Suter-Riniker, Christoph Aebi, Christian Beuret, Daniel H. Paris, Stephen L Leib, Alban Ramette

## Abstract

**Background:** Human Adenoviruses (HAdVs) are highly contagious pathogens of clinical importance, especially among the pediatric population. Studies on comparative viral genomic analysis of cases associated with severe and mild infections due to HAdV are limited. Using whole-genome sequencing (WGS), we investigated whether there were any differences between circulating HAdV strains associated with severe infections (meningitis, sepsis, convulsion, sudden infant death syndrome, death, and hospitalization) and mild clinical presentations in pediatric patients hospitalized between the years 1998 and 2017 in a tertiary care hospital group in Bern, Switzerland covering a population base of approx. 2 million inhabitants. The HAdV species implicated in causing severe infections in this study included HAdV species C genotypes (HAdV1, HAdV2, and HAdV5). Clustering of the HAdV whole-genome sequences of the severe and mild cases did not show any differences except for one sample (isolated from a patient presenting with sepsis, meningitis, and hospitalization) that formed its own cluster with HAdV species C genotypes. This isolate showed intertypic recombination events involving four genotypes, had the highest homology to HAdV89 at complete genome level, but possessed the fiber gene of HAdV1, thereby representing a novel genotype of HAdV species C. The incidence of potential recombination events was higher in severe cases than in mild cases. Our findings confirm that recombination among HAdVs is important for molecular evolution and emergence of new strains. Therefore, further research on HAdVs, particularly among susceptible groups, is needed and continuous surveillance is required for public health preparedness including outbreak investigations.

## Introduction

Human adenoviruses (HAdVs) are pathogens of clinical importance and often used as model to understand biological systems [1]. As public health concern, HAdV infections are common and often cause acute diseases ranging from mild to severe, including death especially in immunocompromised individuals [2-6]. HAdVs are non-enveloped, double-stranded deoxyribonucleic acid (DNA) viruses belonging to the *Adenoviridae* family within the genus *Mastadenovirus*. At present, over 100 genotypes of HAdVs are recognized by the Human Adenovirus Working Group July, 2019 Update (http://hadvwg.gmu.edu/), which are classified into seven species/groups (HAdV-A to HAdV-G) initially defined on the basis of their physical, chemical and biological properties by serum neutralization methods, but more recently distinguished based on whole-genome sequencing (WGS), genomic and bioinformatic analysis (http://hadvwg.gmu.edu/).

The HAdVs that are predominately reported to be associated with human disease globally include genotypes from species C (HAdV1, HAdV2, and HAdV5), species B (HAdV3 and HAdV7), species E (HAdV4), and species F (HAdV41) [7-11]. In the pediatric population, HAdV infections are predominately caused by genotypes of the HAdV species C [10, 12, 13]. Although HAdV species C consists of only 6 genotypes so far (HAdV1, HAdV2, HAdV5, HAdV6, HAdV57 and HAdV89), these are reported to be more clinically significant than other HAdV species in causing severe infections with life-threatening implications in young children and immunocompromised patients [3, 13]. More than half of HAdV infections in young children are associated with HAdV1 and HAdV2 [2, 10]. After primary infection, genotypes of HAdV-C species may establish latent infections and are capable of long-term persistence in lymphoid cells [14-16]. Thus, asymptomatic individuals can shed infectious viruses in stool for many years [17, 18].

HAdV genomes can be unstable as they are subjected to genetic drift resulting from base insertion, substitutions and deletions, but also are prone to changes observed as antigenic shifts originating from genomic recombination between at least two viral strains [19]. Early studies of HAdV recombination [20-22] led to the hypothesis that molecular evolution of HAdV strains may be driven by recombination [19]. Moreover, recent studies have shown that recombination among circulating HAdV strains is frequent and plays a critical role in shaping the phylogenetic relationships among HAdV genomes [23]. To date, no recombinant HAdV has been reported in Switzerland. Typing systems based on the partial gene sequence of one or more of the three major capsid genes (penton base, hexon, and fiber) are sufficient for gaining insight into the epidemiology of circulating HAdV strains. However, as homologous recombination of HAdV capsid genes plays a central role in shaping the evolution of HAdVs [24-26] and may have consequences for HAdV detection and pathogenicity, information obtained on single gene sequences does not provide enough molecular resolution. Therefore, whole-genome sequencing based methods are recommended to investigate adenovirus genetics.

The aim of our study was to determine if there were any genomic differences between circulating HAdV strains causing severe and mild clinical presentations in hospitalized pediatric patients in Bern, Switzerland. We investigated the phylogenomic relationships and potential recombination events among Swiss HAdV isolates based on their whole-genome sequences. Overall, our results document that recombination is a major factor for HAdV evolution and may possibly contribute to disease severity.

## Methods

### Samples

We defined severe cases as those associated with death or requiring hospitalization. HAdV isolates from patients presenting with severe cases and a subset of selected mild cases were obtained from the Institute for Infectious Diseases (IFIK) clinical bio-bank. Selection criteria of HAdV isolates associated with mild cases was based on HAdV hexon sequences that phylogenetically clustered in the same as or different branches than the HAdV severe cases (Figure 1), as determined previously [10]. Patient samples for this study were approved by the Swiss Ethics Committees on Research involving humans (BASEC-Nr:Req-2018-00158).

**Figure 1.**
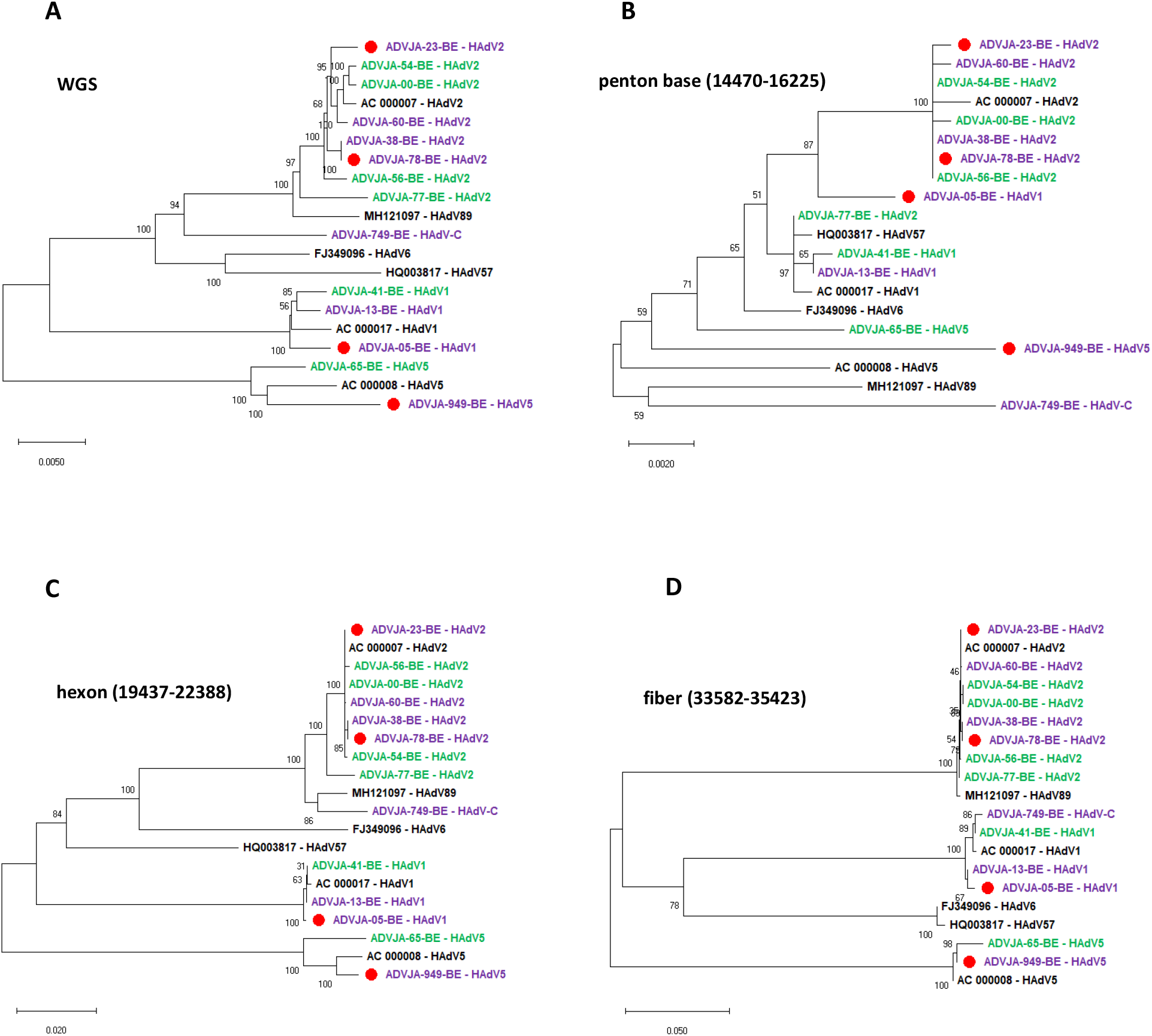
Phylogenetic analysis of HAdV genotypes identified from severe and mild cases in pediatric patients based on complete genome sequences (A), complete penton base sequences (B), complete hexon gene sequences (C), and complete fiber gene sequences (D). Eight severe cases (highlighted in purple, of those who died indicated with a red circle), six mild cases (highlighted in green), and GenBank sequences of the prototype strains are indicated in bold black font. Labelling indicates isolate names or accession number (for prototype strains), followed by HAdV genotype/species classification. Alignment positions relative to prototype sequence AC_000007 are indicated for each major HAdV gene (penton base, hexon, and fiber). The Neighbor-Joining tree was generated based on Kimura 2-parameter model with 1000 bootstrap replicates and branch lengths represent numbers of substitutions per site.

### Cell lines and virus DNA isolation

Cell cultures were performed in A549 cells purchased from the American Type Culture Collection (Manassas, VA, USA) and were maintained in Earle’s minimal essential medium (MEM) (Biochrom GmbH, Germany) supplemented with 2.2 g/l NaHCO_3_ (Biochrom), 1% L-glutamine (Merck, Germany), 1% penicillin (10,000 U/ml) and streptomycin (10,000 μg/ml) (Biochrom), 1% fungizol (CPS Cito Pharma Services, Switzerland), and 1% heat inactivated Fetal Bovine Serum (FBS) (Biochrom) at 37°C and 5% CO_2_. The samples inoculated in A549 cells were incubated at 37°C for 3-7 days or until a cytopathic effect was observed. After cytopathic effect was confirmed, the positive cell culture supernatant was subjected to DNA extraction. DNA viral extraction from 200 µl supernatant of the cell cultured HAdV positive samples was automatically performed using the NUCLISENS easyMAG (bioMérieux, Geneva, Switzerland) extractor, as per manufacturer’s instructions. After DNA extraction, the DNA concentration of each sample was measured using the Qubit dsDNA high sensitivity assay kit on the Qubit 3.0 Fluorometer (ThermoFisher Scientific, Zug, Switzerland) as per manufacturer’s protocol prior to whole genome amplification (WGA).

### Whole-genome amplification (WGA), preparation of sequencing libraries

The Seqplex enhanced DNA amplification kit (Sigma-Aldrich Chemie GmbH, Buchs SG, Switzerland) was used for WGA of the DNA as per manufacture’s protocol. The DNA was sheared to 400bp using the Covaris M220 system (Covaris Ltd, Brighton, United Kingdom) prior to WGA amplification. The Covaris program for shearing was as follows; temperature 20°C, duty factor 20, cycles/burst 200, peak power 50, and time 60 seconds. Libraries were constructed with the Ion plus fragment library kit using the ABI library builder according to the manufacturer’s instructions (ThermoFisher Scientific).

Sequencing barcoded libraries were prepared automatically on the AB Library Builder System with the Ion plus Fragment Library Kit (ThermoFisher Scientific) according to the Ion Xpress Plus and Ion Plus Library preparation (ThermoFisher Scientific). The generated adapter-ligated libraries were subjected to size selection with 0.55× Agencourt AMPure XP magnetic beads (Beckman Coulter, Nyon, Switzerland), followed by measurement of the concentration using Qubit dsDNA High Sensitivity Assay kit (ThermoFisher Scientific) and assessment of the size distribution using the Agilent 2100 Bioanalyzer system with the Agilent High Sensitivity DNA kit (Agilent Technologies AG, Basel, Switzerland). Sample libraries were pooled and loaded automatically on the 530-chip using the Ion Chef instrument according to the manufacturer’s instructions. The loaded chip was then inserted into the Ion S5XL for sequencing with 850 flows using the Ion 530 (400 bp) chip kit.

### Bioinformatic analyses

Raw sequence data in BAM file format were imported into CLC genomics workbench v12.0.3 (QIAGEN, Aarhus, Denmark) and trimmed with the following parameters: Removal of adaptor/ barcode sequences from both ends, discarding of reads with ambiguous bases, base quality below Q30, and read length below 50 nt. The trimmed reads were exported as fasta file from CLC genomics workbench and imported into Geneious Prime 2020.1.2 software (http://www.geneious.com, [27]) for further analysis: The trimmed reads were normalized to 100x coverage using BBNorm (version 38.37) and duplicate reads were removed using Dedupe (version 38.37; k-mer seed length of 31). Reads were subjected to *de novo* assembly using SPAdes assembler (version 3.13.0) using default parameters. As the genotypes of most samples were previously identified based on Sanger sequencing of the partial hexon gene, the resulting contigs for each sample were mapped to HAdV prototype strain sequences of either HAdV1 (AC-000017), HAdV2 (AC-000007) or HAdV5 (AC-000008) to which it belonged using Bowtie2 (version 2.3.0), with “end-to-end” alignment and default parameters. Consensus sequences were produced and visually inspected. Following visual inspection of consensus sequences, gaps indicating missing nucleotides were identified and corrected by Sanger sequencing. Annotation of the consensus sequences was performed based on HAdV reference prototype strain genome annotations, using the Annotate & Predict tool within Geneious prime [27]. The annotations were manually checked and edited. The genome sequences for all 14 isolates were deposited to the European Nucleotide Archive, under project reference PRJEB40708.

### Phylogenetic and computational analysis

Multiple sequence alignments of HAdV sequences from patients along with HAdV reference prototype sequences for construction of phylogenetic trees were performed using MAFFT v7.450 (within Geneious Prime) with the default gap parameters. For detailed comparisons, sequences of prototype strains HAdV1 (AC-000017), HAdV2 (AC-000007), HAdV5 (AC-000008), HAdV6 (FJ349096), HAdV57 (HQ003817) and HAdV89 (MH121097) belonging to group HAdV-C obtained from GenBank were used for phylogenetic and genetic recombination analysis. Phylogenetic analysis was conducted in MEGAX (version 10.0.3, [28]) using the Neighbor-Joining method with 1000 bootstrap replications and the evolutionary distances computed using the Kimura 2-parameter method [29]. Bootscanning analyses to identify potential recombination events were performed using SimPlot Software version 3.5.1 (https://sray.med.som.jhmi.edu/SCRoftware/simplot/) with parameters consisting of window size of 5000 bp, step size of 100 bp, gap stripping, 100 replicates, kimura (2-parameter), and Neighbor-Joining method. Global pairwise genome comparisons were performed using zPicture (https://zpicture.dcode.org/).

## Results

### Genomic characteristics and comparative analysis

The complete genomes of HAdV isolates from pediatric patients presenting with severe cases including meningitis, sepsis, convulsion, sudden infant death syndrome, death, and hospitalization were analyzed alongside those obtained from HAdV pediatric patients presenting with mild cases (**Table 1**). The selection of mild cases for genomic comparative purposes was based on clustering of their partial hexon gene nucleotide sequences on the phylogenetic tree in comparison to partial hexon sequences isolated from severe cases. All patient isolates in this study were initially typed by Sanger sequencing of the hypervariable hexon region 1-7 [10]. Molecular typing based on this region of a diagnostic specimen is sufficient to identify circulating HAdV genotypes causing infection among hospitalized patients. However, it does not allow for precise and accurate resolution among genotypes in particular identifying and assessing potential recombination events along with genome rearrangements. The HAdV genotypes analyzed in this study belonged to species C as these were the ones implicated in causing severe infection including death among the pediatric patients in our study collection. Of the initial 14 HAdV species C samples analyzed, three belonged to HAdV1, nine belonged to HAdV2 and two belonged to HAdV5 (**Table 1**). The genome length of samples identified as HAdV1 ranged from 35,858 bp to 35974 bp, HAdV2 from 35,813 bp to 35,951 bp, and HAdV5 from 35,872 bp to 35,881 bp. The G+C content of genotypes belonging to HAdV species C (i.e., HAdV1, HAdV2 and HAdV5) varied between 55.20 % and 55.40%. The percentage nucleotide identities between the complete genome sequences of the HAdV prototypes and strains of each genotype identified in this study was 97.58 – 98.97% among HAdV1 strains, 98.84 – 99.4% among HAdV2 strains, and 98.11 – 98.38% among HAdV5 strains. A slight variation in the percentage similarity at the nucleotide and amino acid level based on the three major capsid genes (penton-base, hexon, and fiber) of the HAdV1, HAdV2 and HAdV5 strains in this study was observed (**Table 2**).

**Table 1.** Characteristics of HAdV isolates in this study.

| Isolate | Genotype (partial hexon) | Year of isolation | Patient's age range | Patient's gender | Specimen type | Clinical symptoms and outcome* | Genome size (bp) | Genotype classification (WGS) |
| --- | --- | --- | --- | --- | --- | --- | --- | --- |
| ADVJA-13-BE | HAdV1 | 2002 | 1-5 y | F | upper respiratory tract | **meningitis** | 35,858 | HAdV1 |
| ADVJA-05-BE | HAdV1 | 2014 | 1-5 y | M | lower respiratory tract | **sudden infant death syndrome** | 35,974 | HAdV1 |
| ADVJA-41-BE | HAdV1 | 2014 | 1-5 y | M | stool | gastroenteritis | 35,859 | HAdV1 |
| ADVJA-60-BE | HAdV2 | 2000 | < 1 y | M | stool | **gastroenteritis, hospitalization** | 35,839 | HAdV2 |
| ADVJA-749-BE | HAdV2 | 2002 | 1-5 y | M | upper respiratory tract | **sepsis, meningitis, hospitalization** | 35,853 | HAdV-C |
| ADVJA-23-BE | HAdV2 | 2005 | 1-5 y | M | lower respiratory tract | **death** | 35,813 | HAdV2 |
| ADVJA-78-BE | HAdV2 | 2006 | 1-5 y | M | lower respiratory tract | **sudden infant death syndrome** | 35,890 | HAdV2 |
| ADVJA-00-BE | HAdV2 | 2006 | 1-5 y | F | upper respiratory tract | meningitis suspected, but not confirmed; no hospitalization | 35,843 | HAdV2 |
| ADVJA-56-BE | HAdV2 | 2006 | 1-5 y | F | stool | gastroenteritis | 35,884 | HAdV2 |
| ADVJA-54-BE | HAdV2 | 2006 | 1-5 y | F | stool | abdominal pain, fever, gastroenteritis, no therapy | 35,826 | HAdV2 |
| ADVJA-77-BE | HAdV2 | 2009 | < 1 y | M | upper respiratory tract | cough, upper respiratory infection | 35,951 | HAdV2 |
| ADVJA-38-BE | HAdV2 | 2010 | < 1 y | M | upper respiratory tract | **fever, convulsions, hospitalization** | 35,883 | HAdV2 |
| ADVJA-949-BE | HAdV5 | 2012 | < 1 y | F | lower respiratory tract | **sudden infant death syndrome** | 35,872 | HAdV5 |
| ADVJA-65-BE | HAdV5 | 2013 | 1-5 y | F | stool | oncological immunosuppressed, gastroenteritis | 35,881 | HAdV5 |

\*Clinical symptoms indicated in bold are severe cases of HAdV infection.

**Table 2.** Percentage (%) similarity at nucleotide and amino acid level of HAdV1, HAdV2, and HAdV5 strains in this study. The penton base, hexon and fiber were chosen because they play an important role in determining tissue tropism.

| **Region** | **% similarity at** | **HAdV1**<br>**(N=3)** | **HAdV2**<br>**(N=9)** | **HAdV5**<br>**(N=2)** |
| --- | --- | --- | --- | --- |
| **Penton** | nucleotide | 98.90 - 99.94 | 99.36 - 100 | 97.80 |
|  | amino acid | 98.61 - 99.83 | 99.47 - 100 | 98.26 |
| **Hexon** | nucleotide | 99.79 - 99.97 | 98.69 - 100 | 96.75 |
|  | amino acid | 100 | 99.79 - 100 | 98.74 |
| **Fiber** | nucleotide | 99.20 - 99.66 | 99.71 - 100 | 98.17 |
|  | amino acid | 98.80 - 99.14 | 99.66 - 100 | 97.76 |

### Phylogenetic analysis

Phylogenetic analysis was performed on whole-genome sequences and on complete sequences of penton base, hexon and fiber genes to investigate the genetic relationships between the Swiss HAdV strains and the prototype HAdV strains obtained from GenBank (**Figure 1**). Whole-genome phylogenetic analysis of the eight severe and six mild cases did not show any differences except for ADVJA-749-BE (isolated from a patient presenting with sepsis, meningitis and was hospitalized) that formed a separate cluster among other HAdV species C genotypes (**Figure 1A**). Overall, clustering of the complete hexon and fiber gene sequences agreed with clustering of the complete genomic sequences except for isolate ADVJA-749-BE with intertypic recombination affecting the hexon and fiber gene (**Figures 1A, 1C, and 1D**). Phylogenetic analyses of the penton base gene showed a completely different clustering with relatively low bootstrap support values (**Figure 1B**). This suggests high identity within the penton base gene between genotypes of the same HAdV species. Phylogenetic analysis of the three HAdV coding regions (penton base, hexon, and fiber) demonstrated that ADVJA-749-BE strain exhibited a close relationship to HAdV89 in its penton base, and hexon gene, and to HAdV1 in its fiber gene (**Figure 1B, 1C, and 1D**, respectively).

### Comparative genomic analyses of a potentially “novel” HAdV-C genotype

To determine the genomic characterization of ADVJA-749-BE strain, comparison with all prototype strains of HAdV species C was performed. Compared with the complete HAdV species C sequences of the 6 prototype strains of HAdV1 (AC_000017.1), HAdV2 (AC_000007.1), HAdV5 (AC_000008), HAdV6 (FJ349096), HAdV57 (HQ003817), and HAdV89 (MH121097), ADVJA-749-BE strain shares the highest nucleotide similarity (97.37%) at the full genome level with the prototype strain HAdV89 (**Table 3**). Highest similarity with this prototype strain was also observed within the penton base (98.31%), hexon (98.01%), E1B 55K (99.73%) and the E3 ORF (99.58%). The latter region of ADVJA-749-BE also had the same nucleotide identity (99.58%) with the prototype strain of HAdV2 and HAdV6. The prototype strains HAdV2 and HAdV6 also showed the greatest similarities to ADVJA-749-BE in the E1A (99.89%). Furthermore, prototype strain HAdV2 showed the highest similarities with ADVJA-749-BE within the DBP (99.06%) and 100K (99.63%) genes. Within the DNA polymerase gene, ADVJA-749-BE shared highest similarity (99.67%) to HAdV6. On the other hand, the ADVJA-749-BE strain shared higher identities with HAdV1 strain in the fiber region (**Table 3**). A high level of similarity and identity across the genomes of HAdV species C was observed (**Figure 2**). The results obtained complement the phylogenetic analysis demonstrating that the ADVJA-749-BE strain is highly similar to the prototype strains of HAdV species C but with differences across the entire genome particularly showing diversity in the hexon, E3 and fiber (**Figure 2**).

**Table 3.** Percentage nucleotide (nt) sequence identities between ADVJA-749-BE and representative prototype HAdV species C genotypes.

| Region | % nt identity |  |  |  |  |  |
| --- | --- | --- | --- | --- | --- | --- |
|  | ADVJA-749-BE strain |  |  |  |  |  |
|  | HAdV1 | HAdV2 | HAdV5 | HAdV6 | HAdV57 | HAdV89 |
| E1A | 98.28% | **99.89%** | 99.31% | **99.89%** | 99.77% | 99.20% |
| E1B 55k | 97.80% | 99.66% | 97.93% | 99.60% | 98.00% | **99.73%** |
| DNA polymerase | 98.64% | 99.30% | 99.30% | **99.67%** | 98.64% | 99.33% |
| Penton base | 97.33% | 97.73% | 98.14% | 97.45% | 97.33% | **98.31%** |
| Hexon | 85.12% | 97.39% | 81.98% | 87.55% | 86.76% | **98.01%** |
| DBP | 96.60% | **99.06%** | 97.30% | 98.81% | 97.36% | 97.55% |
| 100K | 98.23% | **99.63%** | 96.83% | 99.38% | 97.98% | 98.64% |
| Fiber | **99.31%** | 71.95% | 74.60% | 70.38% | 70.27% | 71.89% |
| E3 ORF | 87.71% | **99.58%** | 86.04% | **99.58%** | 97.29% | **99.58%** |
| E4 ORF | 99.55% | **99.77%** | 99.32% | 99.10% | 97.74% | 98.87% |
| Full genome | 95.29% | 97.31% | 93.54 | 96.37% | 95.50% | **97.37%** |

**Figure 2.**
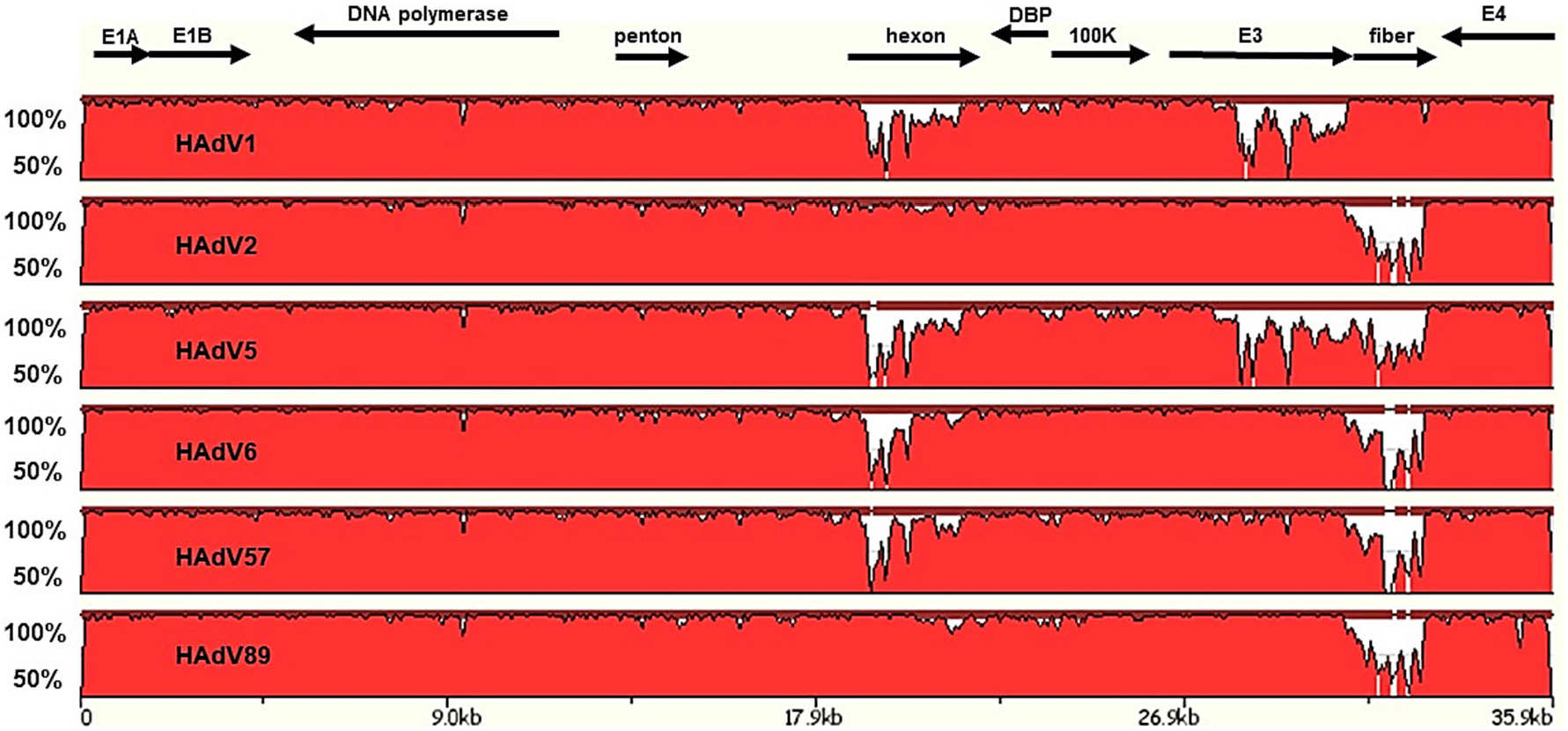
Pairwise genome comparative analysis of HAdV species C strains. zPicture analysis based on BlastZ local alignment algorithm to show the regions of nucleotide sequence similarity between ADVJA-749-BE strain (used as query) and the representative prototype strains of HAdV species C. The Y-scale represents the percentage identity of genome pairs from 50% and 100%. Arrows indicate approximate positions of the coding transcripts and their orientation.

### Genomic recombination analysis

To investigate potential recombination events within and between HAdV genomes isolated in this study, particularly the genome sequences isolated from HAdV severe cases, we performed Bootscan analysis with studied Swiss strains and representative HAdV species C prototype strains available in GenBank (**Figure 3**). Of note was isolate ADVJA-749-BE which was associated with multiple recombination events. The genome sequence of ADVJA-749-BE that was isolated from a pediatric patient presenting with sepsis, meningitis and hospitalized was identified as a possible recombinant that may have arisen from recombination events involving HAdV1, HAdV2, HAdV5, and HAdV89. The other samples from severe and mild cases also showed some potential recombination events but to a lesser extent (**Figure 3, S1**). For severe infections due to HAdV species C, 75% (6 out of 8 cases) had potential recombination events involving at least two of the representative HAdV species C prototype strains, while 66.7% (4 out of 6 cases) of the mild infections had potential recombination events involving also at least two of the HAdV species C prototype strains.

**Figure 3.**
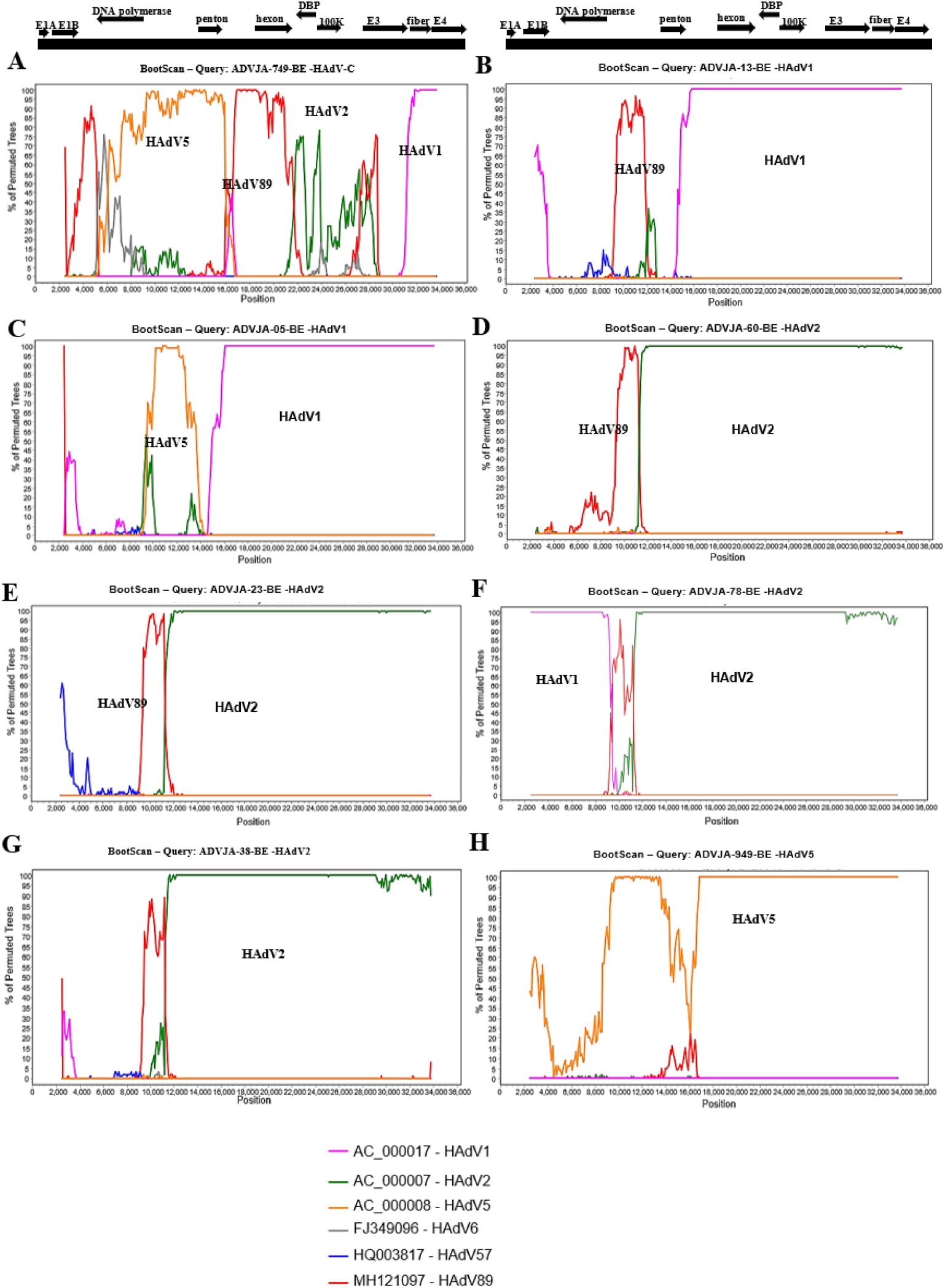
Bootscan analysis of the whole-genome sequences for severe HAdV isolates compared with sequences of prototype HAdV1, HAdV2, HAdV5, HAdV6, HAdV57, and HAdV89. Bootscan of whole-genome sequence from patient presenting with sepsis, meningitis and was hospitalized due to HAdV species C genotype (A), meningitis due to HAdV1 (B), sudden infant death syndrome due to HAdV1 (C), gastroenteritis and hospitalization due to HAdV2 (D), death resulting from HAdV2 (E), sudden infant death syndrome due to HAdV2 (F), fever, convulsion, and hospitalization due to HAdV2 (G), and sudden infant death syndrome due to HAdV5 (H). The genotypes involved in recombination events for each of the severe cases are indicated on each panel. The black bar at the top represents the genome map with black arrows indicating approximate position of the coding transcripts and their direction. The legend shows the representative prototype HAdV species C strains used for comparison with the labelling as accession number – HAdV genotype. The percentage of permutated trees that supported grouping are marked along the y-axis and the genome nucleotide position are indicated along the x-axis. Parameter setting for the recombination analysis using Bootscan in the Simplot software were: window size (5,000 nucleotides), step size (100 nucleotides), replicates used (n =100), gap stripping (on), distance model (Kimura) and tree model (Neighbor-joining).

## Discussion

We conducted comparative genomic analysis of whole-genome sequences of HAdV species C from hospitalized pediatric patients in Bern, Switzerland. Among the six genotypes of HAdV species C, HAdV1 and HAdV2 were previously reported to cause higher morbidity than other genotypes among the pediatric patients hospitalized between the year 1998 and 2017 in a tertiary care hospital group in Bern, Switzerland covering a population base of 2 million inhabitants [10]. A trend toward more severe cases due to HAdV species C has also been reported by Esposito and colleagues [30]. To date, no recombinant HAdV has been reported or identified in Switzerland, mainly because of a lack of studies on HAdV epidemiology and molecular evolution. Consequently, fourteen whole-genome HAdV-C sequences (eight from severe cases and six from mild cases) were generated. The sequences were subjected to phylogenetic analyses and probed for potential recombination events and genome rearrangements, which are recognized as important mechanisms that may influence tissue tropism, and potentially pathogenicity and virulence of novel HAdV pathogens [24, 26, 31]. Global pairwise genome comparisons were also performed (**Figure 2**). Our results demonstrated that the phylogenetic analysis of the complete genomes, hexon and the fiber genes in this study provided similar information regarding clustering of the HAdV strains except for one strain (ADVJA-749-BE) showing intertypic recombination affecting the hexon and fiber genes. The penton base phylogeny, however, failed to provide much meaningful information on HAdV species C strains as the 14 sequences did not show much divergence between each other (**Figure 1B**), a result supported by the findings of Zhang and Huang [32]. Moreover, a study by Robinson and colleagues found that HAdV species C and E have similar penton base genes but show diversity in the hexon, fiber, and E3 ORFs [25], which is also observed in our study (**Figure 2)**. Among members of HAdV species C, recombination events were relatively more frequent among severe cases than mild cases (75% versus 66.7%, respectively) (**Figures 3 and S1**).

Of particular note was the identification of a strain (ADVJA-749-BE) that represents a potentially novel HAdV species C genotype isolated from a child presenting with sepsis, meningitis, and hospitalization, thus may be an etiological agent associated with sepsis and meningitis. Although the initial typing results based on the partial hexon gene identified HAdV2 as the cause of infection, phylogenomic analysis indicated that this strain formed a separate cluster among the other genotypes of HAdV species C (**Figure 1A**), sharing the highest nucleotide identity (97.4% and 97.3%) with HAdV89 and HAdV2, respectively at the genome level (**Table 3**), but possessed fiber gene sequence identical to that of HAdV1 (**Figures 1D, 2, and Table 3**). Moreover, recombination analysis further confirmed that the ADVJA-749-BE strain was a recombinant of HAdV1, HAdV2, HAdV5, and HAdV89 (**Figure 3A**). Novel HAdV pathogens are well-known to arise from recombination events occurring only between HAdV genotypes of the same species and in regions of high sequence homology [20, 21]. We therefore propose that ADVJA-749-BE isolate is a potentially novel genotype of HAdV species C, which may be an etiological agent associated with sepsis. Of the HAdV species C genotypes, HAdV57 (isolated from stool of a healthy child in 2001) [33] and HAdV89 (identified from stool of an immunosuppressed patient in 2015) [24] were both identified as recombinant viruses, with HAdV57 having a similar fiber gene to HAdV6 and harboring a unique hexon distinguished by its loop-2 motif [33], whilst HAdV89 had a novel penton base sequence [24]. The circulation of recombinant HAdV species C strains has also been recently reported [32].

For natural recombination to occur, co-infection is required. Co-infection by two or more genotypes of HAdV species has been documented [34]. A study by Lukashev and colleagues, in which they analyzed sixteen HAdV species C field strains at four genomic regions including the hexon, fiber, polymerase and E1A regions, suggested that recombination is frequent [23]. This finding was also supported by a recent study by Dhingra and colleagues which demonstrated that potential multiple recombination events within the E1 and E4 gene regions are likely to contribute to the evolution of species HAdV-C [24].

Overall, recombination and genomic rearrangements within the three major HAdV capsid genes (penton base, hexon, fiber) that are important determinants of tropism, as well as the E3 region that harbors genes affecting the host immunity after virus infection [35] may contribute to disease severity. Nevertheless, further work is needed to verify factors contributing to HAdV disease severity so as to better understand ways of developing effective preventive and therapeutic measures.

## Data Availability

Genome sequences for all 14 isolates are
deposited to the European Nucleotide Archive, under project reference PRJEB40708

## Supplementary Material

The supplementary file S1: Bootscan analysis of the whole-genome sequences obtained from mild HAdV cases compared with the prototype sequences of HAdV1, HAdV2, HAdV5, HAdV6, HAdV57, and HAdV89.

## Acknowledgements

We would like to thank Nicole Liechti from the Spiez Laboratory for useful discussions.

## References

1. Russell, W.C., Adenoviruses: update on structure and function. J Gen Virol, 2009. 90(Pt 1): p. 1–20.

2. Edwards, K.M., Thompson, J., Paolini, J., Wright, P. F., Adenovirus infections in young children Pediatrics, 1985. 76(3): p. 420–424.

3. Garcia, M., A. Beby-Defaux, and N. Leveque, Respiratory viruses as a cause of sudden death. Expert Rev Anti Infect Ther, 2016. 14(4): p. 359–63.

4. Steiner, I., et al., Fatal adenovirus hepatitis during maintenance therapy for childhood acute lymphoblastic leukemia. Pediatric Blood & Cancer, 2008. 50(3): p. 647–649.

5. Lion, T., Adenovirus infections in immunocompetent and immunocompromised patients. Clin Microbiol Rev, 2014. 27(3): p. 441–62.

6. Davis, D., Henslee, P. J., Markesbery, W. R., Fatal adenovirus meningoencephalitis in a bone marrow transplant patient. Annals of neurology, 1988. 23(4): p. 385–389.

7. Lynch, J.P., 3rd, M. Fishbein, and M. Echavarria, Adenovirus. Semin Respir Crit Care Med, 2011. 32(4): p. 494–511.

8. Yliharsila, M., et al., Genotyping of clinically relevant human adenoviruses by array-in-well hybridization assay. Clin Microbiol Infect, 2013. 19(6): p. 551–7.

9. Barrero, P.R., et al., Molecular typing of adenoviruses in pediatric respiratory infections in Buenos Aires, Argentina (1999-2010). J Clin Virol, 2012. 53(2): p. 145–50.

10. Akello, J.O., et al., Epidemiology of Human Adenoviruses: A 20-Year Retrospective Observational Study in Hospitalized Patients in Bern, Switzerland. Clin Epidemiol, 2020. 12: p. 353–366.

11. Guo, L., et al., Detection of three human adenovirus species in adults with acute respiratory infection in China. Eur J Clin Microbiol Infect Dis, 2012. 31(6): p. 1051–8.

12. Tsoumakas, K., et al., Epidemiology of viral infections among children undergoing hematopoietic stem cell transplant: Alpha prospective single-center study. Transpl Infect Dis, 2019. 21(4): p. e13095.

13. Wurzel, D.F., et al., Adenovirus species C is associated with chronic suppurative lung diseases in children. Clin Infect Dis, 2014. 59(1): p. 34–40.

14. Garnett, C.T., et al., Latent species C adenoviruses in human tonsil tissues. J Virol, 2009. 83(6): p. 2417–28.

15. Kosulin, K., et al., Persistence and reactivation of human adenoviruses in the gastrointestinal tract. Clin Microbiol Infect, 2016. 22(4): p. 381 e1–381 e8.

16. Garnett, C.T., et al., Prevalence and quantitation of species C adenovirus DNA in human mucosal lymphocytes. Journal of virology, 2002. 76: p. 10608–10616.

17. Wold, W. and M. Horwitz, Adenoviruses In: Knipe DM, Howley PM, editors. Fields Virology.2007, Philadelphia (PA): Lippincott Williams & Wilkins.

18. Adrian, T., et al., Persistent enteral infections with adenovirus types 1 and 2 in infants. Epidemiol. Infect., 1988. 101: p. 503–509.

19. Crawford-Miksza, L.K., Schnurr, D. P., Adenovirus serotype evolution is driven by illegitimate recombination in the hypervariable regions of the hexon protein. Virology, 1996. 224(2): p. 357–367.

20. Williams, J., et al., Adenovirus recombination: physical mapping of crossover events.. Cell, 1975. 4(2): p. 113–119.

21. Boursnell, M.E., Mautner, V., Recombination in adenovirus: crossover sites in intertypic recombinants are located in regions of homology. Virology, 1981. 112(1): p. 198–209.

22. Mautner, V., Mackay, N., Recombination in adenovirus: analysis of crossover sites in intertypic overlap recombinants. Virology, 1984. 139(1): p. 43–52.

23. Lukashev, A.N., et al., Evidence of frequent recombination among human adenoviruses. J Gen Virol, 2008. 89(Pt 2): p. 380–388.

24. Dhingra, A., et al., Molecular Evolution of Human Adenovirus (HAdV) Species C. Sci Rep, 2019. 9(1): p. 1039.

25. Robinson, C.M., et al., Molecular evolution of human species D adenoviruses. Infect Genet Evol, 2011. 11(6): p. 1208–17.

26. Walsh, M.P., et al., Evidence of molecular evolution driven by recombination events influencing tropism in a novel human adenovirus that causes epidemic keratoconjunctivitis. PLoS One, 2009. 4(6): p. e5635.

27. Kearse, M., et al., Geneious Basic: an integrated and extendable desktop software platform for the organization and analysis of sequence data. Bioinformatics, 2012. 28(12): p. 1647–9.

28. Kumar, S., et al., MEGA X: Molecular Evolutionary Genetics Analysis across Computing Platforms. Mol Biol Evol, 2018. 35(6): p. 1547–1549.

29. Kimura, M., A simple method for estimating evolutionary rates of base substitutions through comparative studies of nucleotide sequences. J Mol Evol, 1980. 16(2): p. 111–20.

30. Esposito, S., et al., Epidemiology and Clinical Characteristics of Respiratory Infections Due to Adenovirus in Children Living in Milan, Italy, during 2013 and 2014. PLoS One, 2016. 11(4): p. e0152375.

31. Dehghan, S., et al., Computational analysis of four human adenovirus type 4 genomes reveals molecular evolution through two interspecies recombination events. Virology, 2013. 443(2): p. 197–207.

32. Zhang, W. and L. Huang, Genome Analysis of A Novel Recombinant Human Adenovirus Type 1 in China. Sci Rep, 2019. 9(1): p. 4298.

33. Walsh, M.P., et al., Computational Analysis of Two Species C Human Adenoviruses Provides Evidence of a Novel Virus. Journal of Clinical Microbiology, 2011. 49(10): p. 3482–3490.

34. Wang, S.L., et al., High-incidence of human adenoviral co-infections in taiwan. PLoS One, 2013. 8(9): p. e75208.

35. Lichtenstein, D.L., Toth, K., Doronin, K., Tollefson, A. E., Wold, W. S., Functions and mechanisms of action of the adenovirus E3 proteins. International reviews of immunology, 2004. 23(1-2): p. 75–111.

