## Supplemental Figure S1 for "Genomic analyses of human adenoviruses unravel novel recombinant genotypes associated with severe infections in pediatric patients"

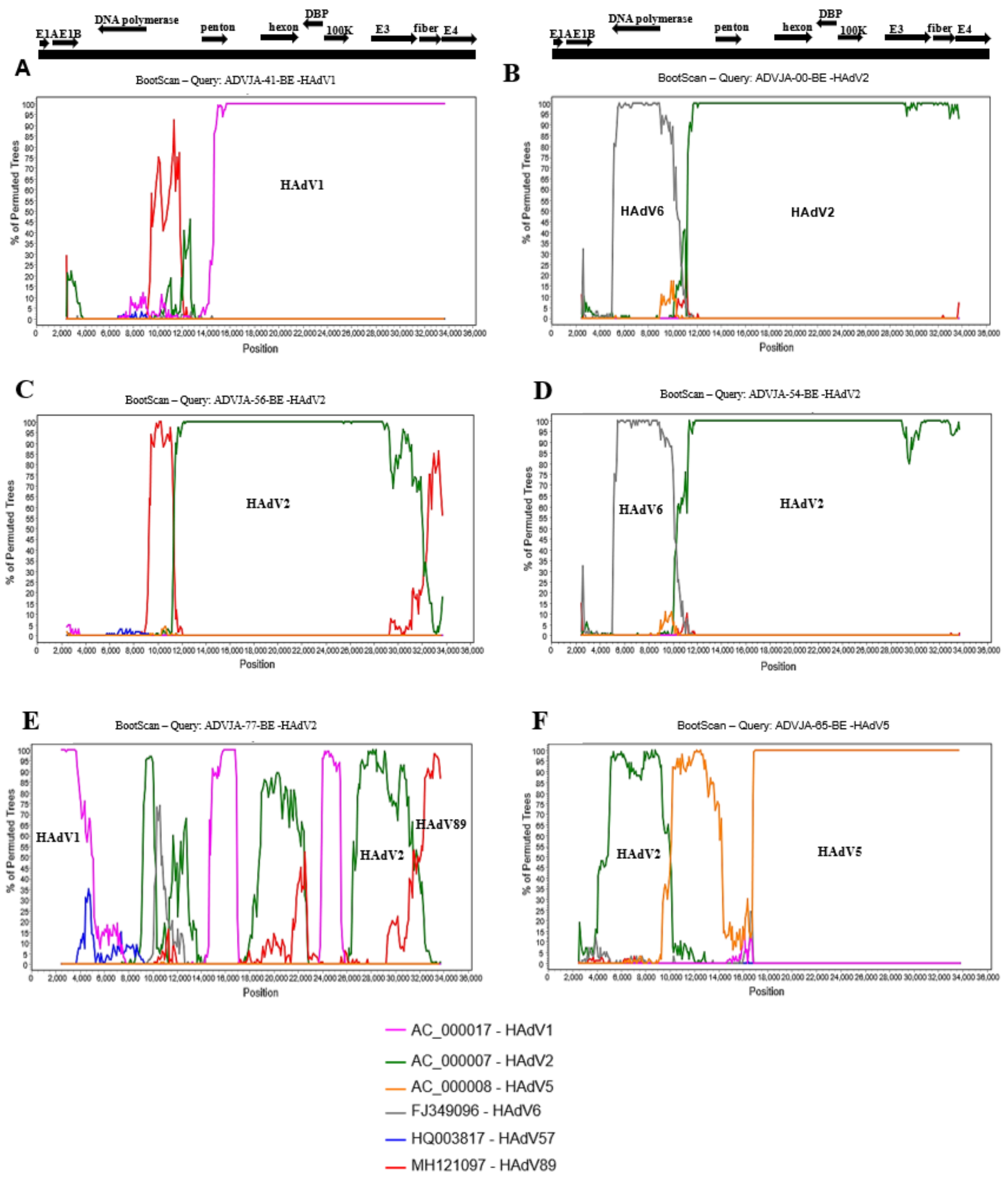


Figure S1: Bootscan analysis of the whole-genome sequences obtained from mild HAdV cases compared with the prototype sequences of HAdV1, HAdV2, HAdV5, HAdV6, HAdV57, and HAdV89. Bootscan of whole-genome sequence from patient presenting with gastroenteritis due to HAdV1 (A), meningitis suspected but not confirmed and no hospitalization due to HAdV2 (B), gastroenteritis due to HAdV2 (C), abdominal pain, fever, gastroenteritis and no therapy due to HAdV2 (D), cough and upper respiratory infection due to HAdV2 (E), oncological, immunosuppressed, and gastroenteritis due to HAdV5 (F).The genotypes involved in recombination events for each of the mild cases are indicated on each panel. The black bar at the top represents the genome map with the black arrows indicating approximate position of the coding transcripts and their direction. The legend shows the representative prototype HAdV species C strains used for comparison with the labelling as accession number – HAdV genotype. The percentage of permutated trees that supported grouping are marked along the y-axis and the genome nucleotide position are indicated along the x-axis. Parameter setting for the recombination analysis using Bootscan in the Simplot software were: window size (5000 nucleotides), step size (100 nucleotides), replicates used (n =100), gap stripping (on), distance model (Kimura) and tree model (Neighbor-joining).
